# Herbal-based medical device efficiently alleviates the symptoms of temporary gastrointestinal diseases - A prospective comparative study

**DOI:** 10.1101/2025.02.25.24317785

**Authors:** Julia Chevts, Nicole Schuster, Kirsten Sander

**Affiliations:** Siteworks Zentrum für klinische Studien Karlsruhe, Karlsruhe, Germany; Retterspitz GmbH & Co. KG, Schwaig bei Nürnberg, Germany; Artimed Medical Consulting GmbH, Kassel, Germany

**Keywords:** gastrointestinal disorders, stomach pain, alternative medicine, flatulence, herbs, medicinal product, heartburn

## Abstract

Gastrointestinal disorders, especially stomach problems, are widespread. This prospective, non-interventional, comparative study investigated safety and efficacy of an herbal medical device for the treatment of stomach disorders. For the first 14 days, the patients did not take the test product and started the intake on day 15 for a duration of 14 days. Full data sets of 100 included patients were assessed. Symptoms were assessed by a visual scale weekly (day 1 to day 28) and changes of the overall recognised symptoms on a 3-stage scale in phase II (day 15 to day 21). Over the whole study duration, the patients documented the daily overall symptom severity in a patient’s diary and all adverse events were assessed. During phase I, most of the symptoms remained unchanged. After phase II, the overall mean severity of gastrointestinal disorders as well as the specific symptom severity had improved significantly. The course of symptom severity was significantly different between both phases, in favour of the study product. Additionally, an average of 56% of patients reported a subjectively reduced symptom severity after day 28. The study product has shown a good efficacy and safety profile in the treatment of gastrointestinal disorders.

## Introduction

The symptoms of gastrointestinal disorders are broad and they often appear without apparent biochemical or organic causes. Affected patients may suffer from pain in the upper and lower abdomen, frequently accompanied with symptoms of heartburn, feeling of abdominal fullness, bloating, discomfort in the stomach, or digestive disorders. These symptoms can be summarized as functional gastrointestinal disorders (FGIDs) ^1^. The severity of FGIDs can be highly variable and patients often consult healthcare professionals because their health-related quality of life is markedly impaired by one or more symptoms ^2^ . The causes of FGIDs are often diverse and heterogenic, including abnormal motility, visceral hypersensitivity, microscopic inflammation, disorders of the brain-gut interaction, psycho-social factors, genetic susceptibility and post-infectious, neuromuscular and neurotransmitter dysfunction ^3^. More than 40% of the people worldwide suffer from at least one FGID, with 49% of women and 37% of men affected ^2, 4^. In Germany, the diagnosis gastrointestinal disorders has a prevalence of 15% and the affected patients mainly suffer from functional dyspepsia (10-40%) or functional irritable bowel syndrome (15-22%) ^5^. With a prevalence of 10-25%, functional abdominal pain is also present in children and adolescents ^6^. The high prevalence as well as the various symptoms and difficulties in diagnosis are an emerging problem in the gastroenterology. The severity of the symptoms can range from feeling discomfort to a markedly lowered quality of life. If FGIDs become chronic, severe inflammation and gastrointestinal diseases may evoke pathological damage to the gastrointestinal tract. More than 50% of patients who consult a healthcare professional due to gastrointestinal disorders received the diagnosis “medically unexplained” after detailed investigations ^2, 7^. Furthermore, co-morbidities like depression, anxiety disorder, and somatic illness can be present and complicate the diagnosis ^5^. More and more patients consult alternative practitioners to find non-conventional sources to treat different gastrointestinal disorders and complementary and alternative medicine (CAM) are increasing in popularity since the nineties ^8, 9^. In 2018, the prevalence of CAM for the treatment of gastrointestinal disorders was reported to be up to 44% in Australian and up to 58% in American adults ^10^. Due to high tolerability and efficacy CAM is the preferred initial therapy by affected patients at present. Many products are over-the-counter medications which can be used by the patients in self-medication. In Germany the rate of using over-the-counter medication increased, especially for indications like coughs and sneezes, pain, stomach and digestive problems ^11^.

Today, several treatment options consist of one or more compounds extracted from plants ^7^. CAM therapies for FGIDs often include peppermint oil, Iberogast (STW 5), Turmeric, Cannabis, Aloe Vera, or Ginger ^6, 9^. For FGIDs, an efficient therapy on herbal basis, which is equivalent to prokinetic drugs, was found in STW 5 for example, a commercial compound of nine herbs ^12^. It has been shown to be efficient in functional dyspepsia and irritable bowel syndrome with a favourable safety profile in several randomized controlled trials ^7, 13^ Nevertheless, due to the high complexity of FGIDs the clinical evidence could only support the use of CAM for a few diseases like dyspepsia and irritable bowel syndrome, physicians still recommend CAM therapies for other mild symptoms. Cognitive behavioural therapy and gut-directed hypnotherapy are further evidenced practices for the treatment of FGIDs, especially irritable bowel syndrome ^5, 9^. If gastro-oesophageal reflux disease (GORD), oesophagitis, dyspepsia, functional dyspepsia, or peptic ulcer are diagnosed, a treatment with antacids, antacid-alginate combinations, H2 receptor antagonists or proton pump inhibitors may be used ^14^. Often the symptoms of patients with FGIDs were managed by Proton-pump inhibitors (PPI) which are known to reduce gastric acidity, but causes a subsequent hypochlorhydia. Patients with FGIDs have a higher mucosal bacterial load in the proximal small bowel, which leads to higher symptom burden and meal-related symptom responses. A treatment with PPI in FGID patients may further increase the bacterial load in the proximal small intestine, which might lead to severe consequences ^15^. FGIDs commonly occur with comorbidities including depression, anxiety disorders, and somatic illness. For example, of patients suffering from irritable bowel syndrome, 15-48% are reported to have somatic illness, 20-70% have depression, and 20-50% have anxiety disorders. The therapeutic drug management of those comorbidities may also induce side effects on the gastrointestinal tract^16^.

Due to the complexity and diversity of treatments, there is still a lack of evidenced data for some products. In this prospective, non-interventional study the safety and efficacy of a further possible CAM therapy for the treatment of FGIDs was investigated. The herbal-based pharmacy-only medical product *Retterspitz Innerlich*^*®*^ is indicated for temporary and chronic gastrointestinal disorders including heartburn, dyspepsia, acidosis/alkalosis, bloating, feeling of fullness and abdominal pain.

## Materials and Methods

### Patients

At three study centres, the recruitment of a prospectively calculated sample size of 100 patients in total was planned. During the outpatient consultation male and female patients between 18 and 85 years of age were recruited who suffered from temporary, gastrointestinal disorders depicted by at least one of the following symptoms: heartburn, dyspepsia, acidosis/alkalosis of the stomach, bloating, feeling of fullness, or abdominal pain. Patients with the following characteristics were excluded: patients with the need of a sustained therapy; patients with renal insufficiency; patients under 18 or over 85 years of age; patients with allergies or incompatibilities to at least one of the ingredients of the study product; pregnancy and/or lactation; incapacity to take part in the study.

### Study design and settings

This study is a two-phase, prospective, non-interventional, national, multicentre study. The objective of this study was to assess the effect of the study product *Retterspitz Innerlich*^*®*^ on the symptoms of gastrointestinal disorders, including heartburn, dyspepsia, acidosis/alkalosis of the stomach, bloating, feeling of fullness, or abdominal pain. For complete datasets, changes of the symptom severity with and without the intake of the study product were analysed over the study period and the development was compared between the groups. Furthermore, the safety of the study product was investigated by documentation and assessment of occurring adverse events (AE) for all included patients during the whole study period of 28 days.

### Study product

*Retterspitz Innerlich*^*®*^ is a medical device and is indicated to balance acid levels in the stomach in case of acute or chronic gastrointestinal problems. It is a milky, nonsterile, colloidal suspension with supplementary aroma of essential oils (orange, lemon, thyme). Adults take the product approximately 15 minutes before having a meal (3-to 5-times daily á 20 ml).

### Study procedure

The study duration was 28 days for each patient and the patients underwent two phases, each lasting 14 days. The initial phase (phase I) covered 14 days without the use of the study product. Symptoms were assessed on day 1, day 7, and day 14. On the last day (day 14) of phase I, the patients received the study product and started with phase II of the study, covering further 14 days. The patients were instructed to take the recommended daily intake of the study product. During phase II, medical gastrointestinal treatments had to be limited to the study product only and other medication for gastrointestinal problems was prohibited. Symptoms were assessed on day 21 and day 28.

During the 28 days study period 5 visits were conducted in total. At each visit, the severity of the symptoms including heartburn, dyspepsia, acidosis/alkalosis of the stomach, bloating, feeling of fullness, or abdominal pain was measured as total burden and symptom specific burden on a 6-stage visual scale (0=no, 1=mild, 2=moderate, 3=strong, 4=very strong, 5=unbelievable strong symptoms). In addition, in phase II, the patients gave a subjective grading of the overall recognised symptoms using a 3-stage scale (0=reduced, 1=remained unchanged, 2=worsened). For a more detailed assessment of the development of the daily impairments caused by symptoms of gastrointestinal disorders, the patients documented the intensity of symptoms in a patient diary on a 6-stage visual scale, which was identical to the scale used at the visits. The assessment of adverse events (AE) and severe adverse events (SAE) of the study product was conducted at every visit. In addition, the documentation of intake behaviour and possible side effects was done by the patients in their patient diaries and discussed with the physicians at each visit.

### Statistics

The statistical analysis was performed with XLSTAT (Vers. 2020.1.1.643772020). All statistical tests were assigned as significant at a significance level of *p=0*.*05* (95% confidence interval). The sample size calculation was performed for two tailed t-test to represent the difference of two independent groups with a power of 0.99.

## Results

110 patients were included in the study to get full data sets of 100 patients. 6 patients had withdrawn their consent, 1 was lost to follow-up, and the other 3 patients had to be excluded from analysis due to incomplete data. Safety data was analysed from all patients, while performance data and general data was only assessed from patients with full data sets. The development of the overall gastrointestinal impairment was analysed based on completed patient diary data of 77 patients. At the baseline visit, besides the existing FGID symptoms, the following comorbidities were reported in 34 patients: reflux oesophagitis (n=17), gastritis (n=10), nausea or vomiting (n=10), diabetes mellitus (n=5), biliary or liver disease (n=3), gastric ulcer (n=1) or pancreatic disease (n=1). Most of the patients suffered from multiple symptoms of gastrointestinal disorders which existed already for at least 14 days. 63% of the patients had 4 or more symptoms concurrently, whereas only 5% of patients showed one single symptom. The distribution of symptoms was as follows: feeling of abdominal fullness (87%), bloating/flatulence (83%), followed by heartburn (69%), irritable stomach (66%), acidosis or alkalosis (54%), and stomach pain (49%). In total, 48% of the patients reported to have treated the symptoms by taking one or more medications; 25% of them have taken proton-pump-inhibitors (PPIs), 22% herbal medications, 16% antacid, 12% simeticone, and 8% other medication. The medication was taken acute complaints (46%) or for more than 3 months (32%). During phase I, a reduced intake was observed. Whereas at day 1, 45 patients reported an intake of at least one medication for the treatment of their gastrointestinal (GI) disorders, at day 7 only 15 patients and on day 14 only 13 patients reported an intake of medication against their GI problems. During Phase II, all patients showed a good compliance and took the study product according to the instruction.

In phase I, changes of the mean severity of the different symptoms were heterogenic. The reported heartburn improved from 1.9±1.5 to 1.8±1.5 (p=0.499), bloating/flatulence from 2.7±1.6 to 2.5±1.4 (p=0.087), acidosis/alkalosis increased from 1.4±1.4 to 1.5±1.5 (p=0.583), and abdominal pain improved from 1.3±1.5 to 1.2±1.4 (p=0.600) from day 1 to day 14 (Fig.1, Fig.2). Significantly improved symptoms were feeling of abdominal fullness (2.7±1.3 to 2.4±1.4; p=0.013) and irritable stomach (1.8±1.5 to 1.6±1.4; p=0.031) (Fig.1, Fig.2). Overall gastrointestinal impairment showed a slightly increasing average severity of the GI disorders from 2.5±1.0 to 2.6±1.3 over time (p=0.297) (Fig.3).

**Figure 1.**
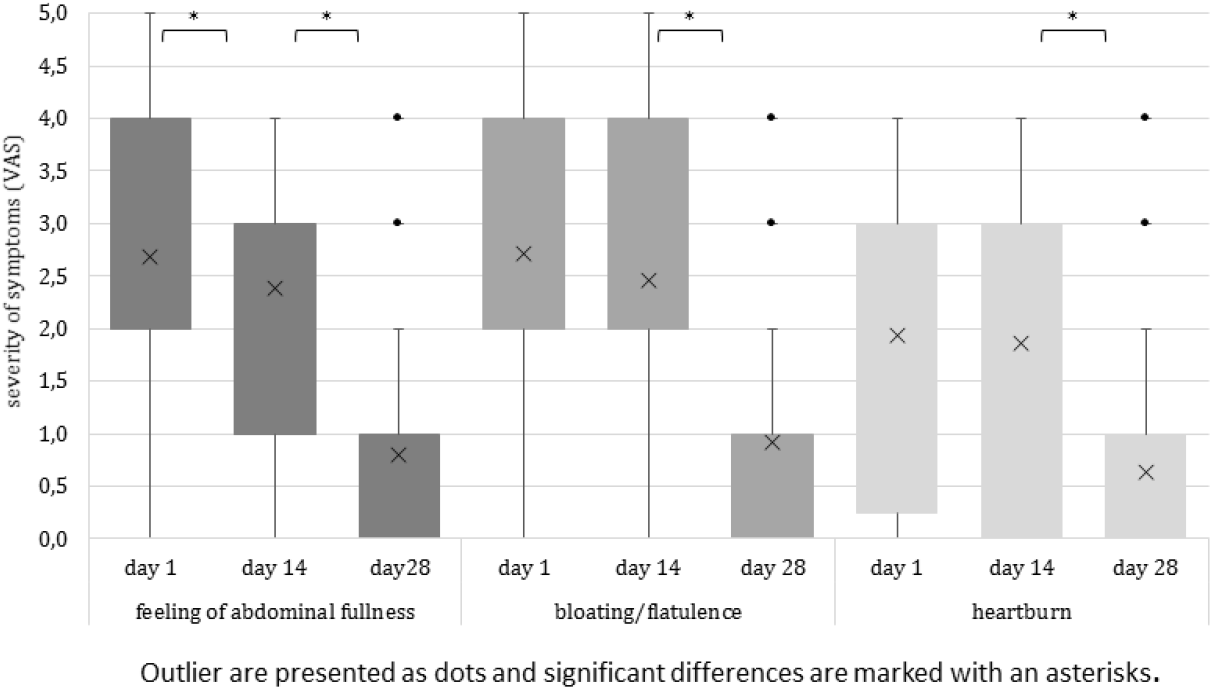
Severity of symptoms

**Figure 2.**
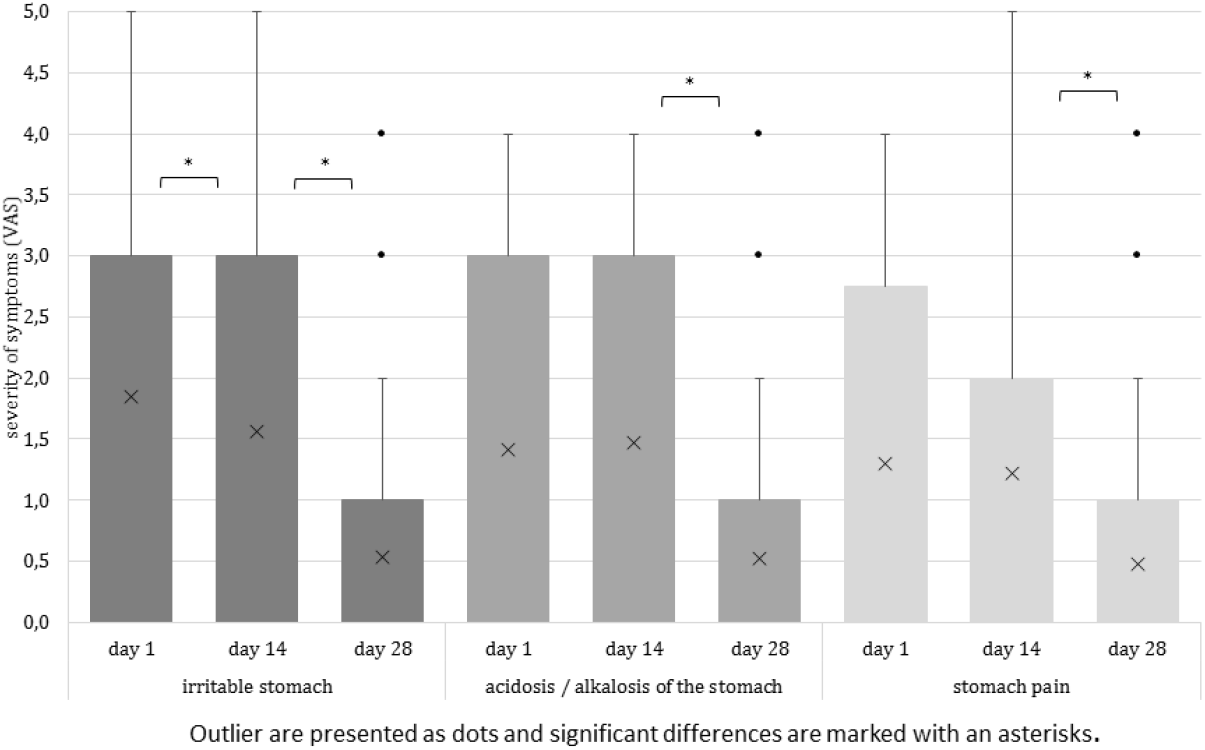
Severity of symptoms

**Figure 3.**
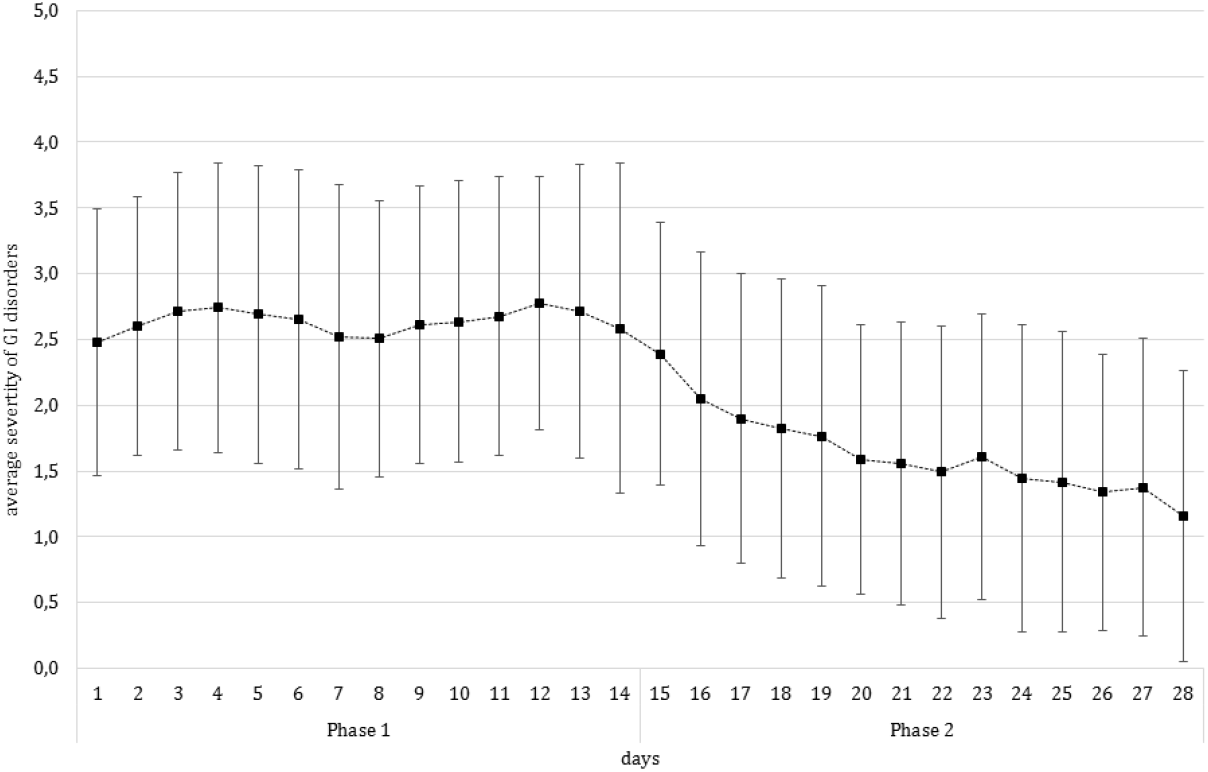
Average severity of GI disorders during Phase 1 and Phase 2 of the study

In phase II, the overall mean severity of the GI disorders was significantly reduced with the intake of the study product from 2.4±1.0 to 1.2±1.1 (p<0.001) (Fig.3). The significant reduction of symptom severity was observed for all different symptoms: feeling of abdominal fullness (from 2.4±1.4 to 0.8±1.), heartburn (from 1.8±1.5 to 0.6±0.9), bloating/flatulence (from 2.5±1.4 to 0.9±1.0), irritable stomach (from 1.6±1.4 to 0.5±0.9), acidosis/alkalosis (from 1.5±1.5 to 0.5±0.9) and stomach pain (from 1.2±1.4 to 0.5±0.9) (all p<0.001) (Fig.1, Fig.2).

The comparison of the course of the overall severity burden was significantly different between both study phases, showing a more or less constant curve in phase I and a slightly fluctuating constant improvement in phase II (p<0.001). Separate analyses of course differences, divided in patients who took medication in phase I and patients who did not, revealed a similar result (p=0.000 for patients with previous medication intake and p<0.0001 for patients without previous medication intake). The assessment of specific symptoms is shown in table 1.

**Table 1:** Severity of symptoms in phase I and phase II for the subgroups with and without medication intake in Phase I.

| Symptoms | Patients who took medication in Phase I | Patients who did not take medication in Phase I |
| --- | --- | --- |
| Heartburn | Phase I: $2.0 \pm 1.4$ to $2.0 \pm 1.3$ ($p = 0.673$)<br>Phase II: $2.0 \pm 1.3$ to $0.7 \pm 0.9$ ($p < 0.0001$) | Phase I: $1.8 \pm 1.5$ to $1.7 \pm 1.6$ ($p = 0.489$)<br>Phase II: $1.7 \pm 1.6$ to $0.6 \pm 1.0$ ($p < 0.0001$) |
| Irritable stomach | Phase I: $2.1 \pm 1.4$ to $1.7 \pm 1.4$ ($p = 0.025$)<br>Phase II: $1.7 \pm 1.4$ to $0.6 \pm 0.9$ ($p < 0.0001$) | Phase I: $1.6 \pm 1.6$ to $1.4 \pm 1.4$ ($p = 0.337$)<br>Phase II: $1.4 \pm 1.4$ to $0.4 \pm 0.8$ ($p < 0.0001$) |
| Abdominal pain | Phase I: $1.7 \pm 1.4$ to $1.4 \pm 1.4$ ($p = 0.165$)<br>Phase II: $1.4 \pm 1.4$ to $0.6 \pm 0.9$ ($p < 0.0001$) | Phase I: $0.9 \pm 1.3$ to $1.0 \pm 1.4$ ($p = 0.527$)<br>Phase II: $1.0 \pm 1.4$ to $0.4 \pm 0.8$ ($p = 0.002$) |
| Acidosis/alkalosis | Phase I: $1.6 \pm 1.5$ to $1.8 \pm 1.5$ ($p = 0.412$)<br>Phase II: $1.8 \pm 1.5$ to $0.6 \pm 0.9$ ($p < 0.0001$) | Phase I: $1.1 \pm 1.4$ to $1.1 \pm 1.4$ ($p = 1.000$)<br>Phase II: $1.1 \pm 1.4$ to $0.4 \pm 0.9$ ($p = 0.000$) |
| Bloating/flatulence | Phase I: $2.5 \pm 1.6$ to $2.3 \pm 1.5$ ($p = 0.361$)<br>Phase II: $2.3 \pm 1.5$ to $0.9 \pm 1.0$ ($p < 0.0001$) | Phase I: $2.9 \pm 1.6$ to $2.6 \pm 1.3$ ($p = 0.104$)<br>Phase II: $2.6 \pm 1.3$ to $0.9 \pm 1.1$ ($p < 0.0001$) |
| Feeling of abdominal fullness | Phase I: 2.7±1.3 to 2.4±1.5 (p=0.52)<br>Phase II: 2.4±1.5 to 0.8±0.9 (p<0.0001) | Phase I: 2.6±1.3 to 2.4±1.3 (p=0.156)<br>Phase II: 2.4±1.3 to 0.8±1.0 (p<0.0001) |

At the visits, the subjective assessment of severity burden after intake of the study product in phase II showed that an average of 50% (after day 21) and 56% (after day 28) of the patients reported a reduced symptom severity. Whereas, 26% (after day 21) and 23% (after day 28) reported no changes, and 2% (after day 21 and day 28) reported worsening of symptoms.

Nearly all patients (99%) reported having regular bowel movement during the whole study period.

Over the whole study period, 17 adverse events occurred in 13 patients. 14 of them were without causal relationship to the study product and two were possibly related (mild and moderate heartburn which did not exist before intake of the study product. One patient mentioned in the patient diary that he had abdominal cramps. The patient did not provide any information on this to the investigator upon request and withdrew study participation. Therefore, severity and causal relation to the study remained unclear. One serious adverse event occurred (uncomplicated sigmoid diverticulitis) which led to hospitalisation, however it was without causal relationship to the study product or study procedure.

## Discussion

The study results show the efficacy and safety of the study product, *Retterspitz Innerlich*^*®*^, for FGIDs including feeling of abdominal fullness, heartburn, bloating/flatulence, irritable stomach, acidosis/alkalosis and stomach pain, independent of the number and type of concurrent symptoms. It was shown that the study product significantly reduces the severity of gastrointestinal disorders. Approximately 50% of patients reported to have reduced GI problems, meaning their subjective well-being improved as well. This is in accordance with the literature, where almost half of the study patients reported their CAM therapy to work well ^18^. Without the study product, only the feeling of abdominal fullness and irritable stomach in the patient collective improved significantly, which are caused by various reasons. The study did not record the eating behaviour of the patients and it cannot be ruled out that they altered the lifestyle behaviour, including their diet. This could have caused changes in their GI impairments. Most of the patients reported regular bowel movements, which would mostly rule out unknown intolerances and allergies, so further investigations in this regard with additional diagnostic parameters would have been beneficial. However, the patients in the study consulted the study centre for the first time regarding their GI disorders and the study displays the routine treatment. CAM therapy is often recommended for mild symptoms ^5, 9^. Patients showed mild to moderate symptoms, so the foremost treatment would be performed with CAM and without invasive diagnostics, hence more detailed parameters are not available for this study.

Most medicine for FGIDs is sold over the counter and easily accessible. In the study, every second patient took at least one of the medications PPI, herbal-based medicine, antacid, simeticone, or other medication to alleviate their symptoms. The positive effect of the study product has been observed in patients that took self-medication as well as in patients that took no medication in phase I. This was true for both, overall impairment and single symptoms. Patients who did not take any medicine in phase I, did not show an improvement of single symptoms. At the end of phase II, all symptoms showed a highly significant improvement. This indicates that *Retterspitz Innerlich*^®^ has a positive effect on patients who did and did not take medicine, but it is unclear how this effect correlates to specific types of medication. The patient collective was too small to allow subgroup analysis and there is little known about the details of the other medicines.

In studies with herbal medicine aiming at improvements for the digestive system, adverse events occurred in 2.4% of patients ^17^. *Retterspitz Innerlich*^®^ is specifically developed to alleviate symptoms of digestive disorders and showed good tolerability, except mild heartburn, which only occurred in 2% of patients. This is in accordance with the data from literature. Additionally, heartburn belongs to the often concurrently occurring symptoms of FGID. Overall, an improvement of symptoms was reported. The study product can therefore be assumed as safe treatment. It can be used in self-medication by patients and leads to positive outcomes, which improves the patients’ quality of life.

### Limitations

The study was not randomised or included placebo controls, but since the outcomes were compared for the same patient collective, possible confounders or bias could be excluded.

## Conclusion

Gastrointestinal disorders can severely affect the quality of life of affected people. The diagnosis of causes often remains unclear. Hence, the current treatment is often multicausal, including alterations of lifestyle with well-balanced nutrition and physical activity, as well as the medication with herbal-based drugs or medical products. In summary, this study with a herbal-based medical product, *Retterspitz Innerlich*^®^ has shown a good safety and efficacy profile.

## Data Availability

All data is available upon reasonable request to the authors.

